# BCG vaccination induces enhanced frequencies of memory T and B cells and dendritic cell subsets in elderly individuals

**DOI:** 10.1101/2020.10.22.20217471

**Authors:** Nathella Pavan Kumar, Chandrasekaran Padmapriyadarsini, Anuradha Rajamanickam, Perumal Kannabiran Bhavani, Arul Nancy, Bharathi Jayadeepa, Nandhini Selveraj, Dinesh Kumar, Rachel Mariam Renji, Vijayalakshmi Venkataramani, Srikanth Tripathy, Subash Babu

## Abstract

**Background:** BCG vaccination is known to induce innate immune memory, which confers protection against heterologous infections. However, the effect of BCG vaccination on the conventional innate and adaptive immune cells subsets is not well characterized.

**Methods:** We investigated the impact of BCG vaccination on the frequencies of T cell, B cell, monocyte and dendritic cell subsets as well as total antibody levels in a group of healthy elderly individuals (age 60-80 years) at one month post vaccination as part of our clinical study to examine the effect of BCG on COVID-19.

**Results:** Our results demonstrate that BCG vaccination induced enhanced frequencies of central and effector memory CD4+ T cells and diminished frequencies of naïve, transitional memory, stem cell memory CD4+ T cells and regulatory T cells. In addition, BCG vaccination induced enhanced frequencies of central, effector and terminal effector memory CD8+ T cells and diminished frequencies of naïve, transitional memory and stem cell memory CD8+T cells. BCG vaccination also induced enhanced frequencies of immature, classical and activated memory B cells and plasma cells and diminished frequencies of naïve and atypical memory B cells. While BCG vaccination did not induce significant alterations in monocytes subsets, it induced increased frequencies of myeloid and plasmacytoid DCs. Finally, BCG vaccination resulted in elevated levels of all antibody isotypes.

**Conclusions:** BCG vaccination was associated with enhanced innate and adaptive memory cell subsets, as well as total antibody levels in elderly individuals, suggesting its potential utility in SARS-Cov2 infection by enhancing heterologous immunity.

## Introduction

Bacillus Calmette-Guerin (BCG) is a live – attenuated vaccine strain of Myobacterium bovis that provides protection against mycobacterial infections such as tuberculosis and leprosy and was first introduced in 1921 (1, 2). In addition to protective immunity to mycobacterial infections, BCG is also known to protect against heterologous infections (the so-called off-target or non – specific effects) (3, 4). Several epidemiological studies have shown a reduction in childhood mortality in BCG vaccinated children as well as lower incidence of respiratory infections (5-9). In addition, randomized controlled trials have shown that BCG vaccination protects against childhood mortality mainly by providing protection against neonatal sepsis and respiratory infections (10-12). The effect of BCG vaccination in protecting against heterologous infections in adults and more specifically, elderly individuals is less well studied.

Two types of immune mechanisms have been postulated to explain this off-target or non – specific effect of BCG against infections. First, BCG is known to induce CD4+ and CD8+ memory T cells in an antigen-independent manner but cytokine-dependent manner, and this process is termed heterologous immunity (13-16). Second, BCG is known to induce a process called trained immunity or innate immune memory in innate cells, especially monocytes and NK cells, such that these cells can respond more actively to secondary or bystander infections (17, 18). However, whether these mechanisms are operational in elderly individuals, who are at higher risk for infections due to waning immunity is not known.

Hence, we examined the induction of T cell, B cell, monocyte, and dendritic cell subsets in response to BCG vaccination in elderly individuals at baseline and one-month post-vaccination along with baseline frequencies in unvaccinated individuals. We also examined the levels of antibody isotypes following vaccination. We demonstrate that BCG vaccination induces significantly enhanced memory T and B cell responses and increased frequencies of DC subsets, as well as total antibody levels, suggesting that BCG can potentially boost immune responses in a non -specific or off-target manner in these elderly individuals.

## Results

### Study population

The demographics of the study population are shown in Table I. From July 2020 through September 2020, 86 individuals were enrolled in the study with 54 in the vaccinated arm and 32 in the unvaccinated arm. All the vaccinated individuals were followed up at month 1 post-vaccination with no loss to follow-up. Median age was 65 (Range: 60-78) years in BCG vaccinated group and 63 years (Range: 60-80) in the unvaccinated group. There were 34 males and 20 females in the BCG vaccinated and 15 males and 17 females in unvaccinated group. In the enrolled population, 26% of BCG vaccinated and 15% of unvaccinated individuals had diabetes mellitus while 15% and 9% had cardiovascular disease respectively. In our cohort 4%-6% were current smokers and there were 6% were alcoholics. Other baseline characteristics were similar between the two arms.

**Table I:**
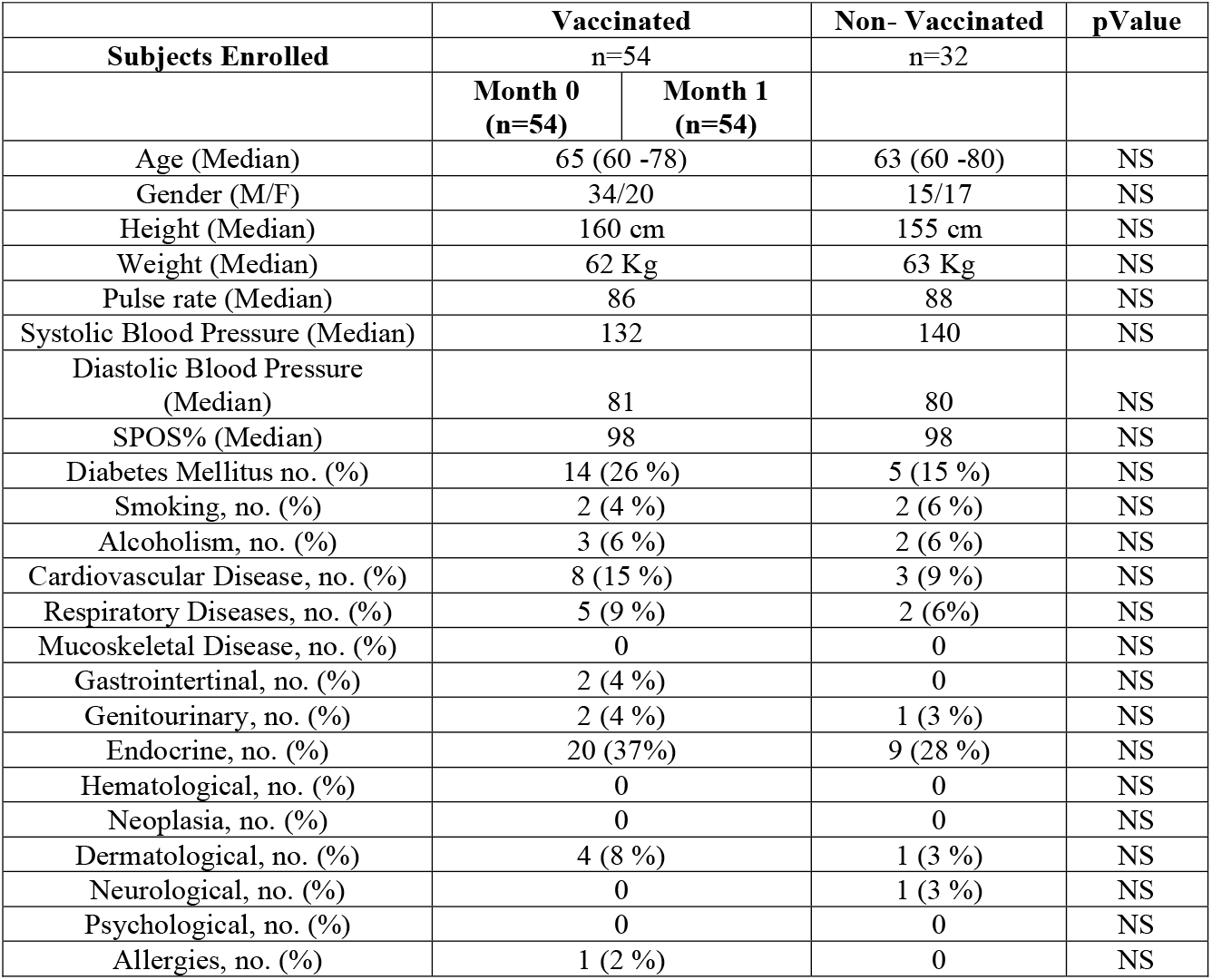
Demographics of the study population.

### BCG vaccination induces enhanced frequencies of central and effector memory CD4+ T cells and diminished frequencies of naïve, transitional, and stem cell memory CD4+ T cells

To assess the ex vivo phenotype of CD4+ T cell subsets following BCG vaccination, we compared the subsets at baseline or before BCG vaccination (M0) and at month 1 (M1) post-vaccination. As shown in Figure 1A, the frequencies of central and effector memory CD4+ T cell subsets were significantly enhanced and the frequencies of naïve, transitional, and stem cell memory CD4+ T cells and regulatory CD4+ T cells were significantly diminished at M1 compared to M0. Next, we compared the frequencies of CD4+ T cell subsets in post-vaccinated individuals to unvaccinated controls. As shown in Figure 1B, BCG vaccinated individuals exhibited increased frequencies of only central memory CD4+ T cells and decreased frequencies of naïve, effector memory, transitional memory, stem cell memory, and regulatory CD4+ T cells. A representative flow cytometry plot showing the gating strategy for CD4+ T cell subsets is shown in S. Figure 1. Thus, BCG vaccination induces enhanced frequencies of central and effector memory CD4+ T cells in elderly individuals.

**Figure 1.**
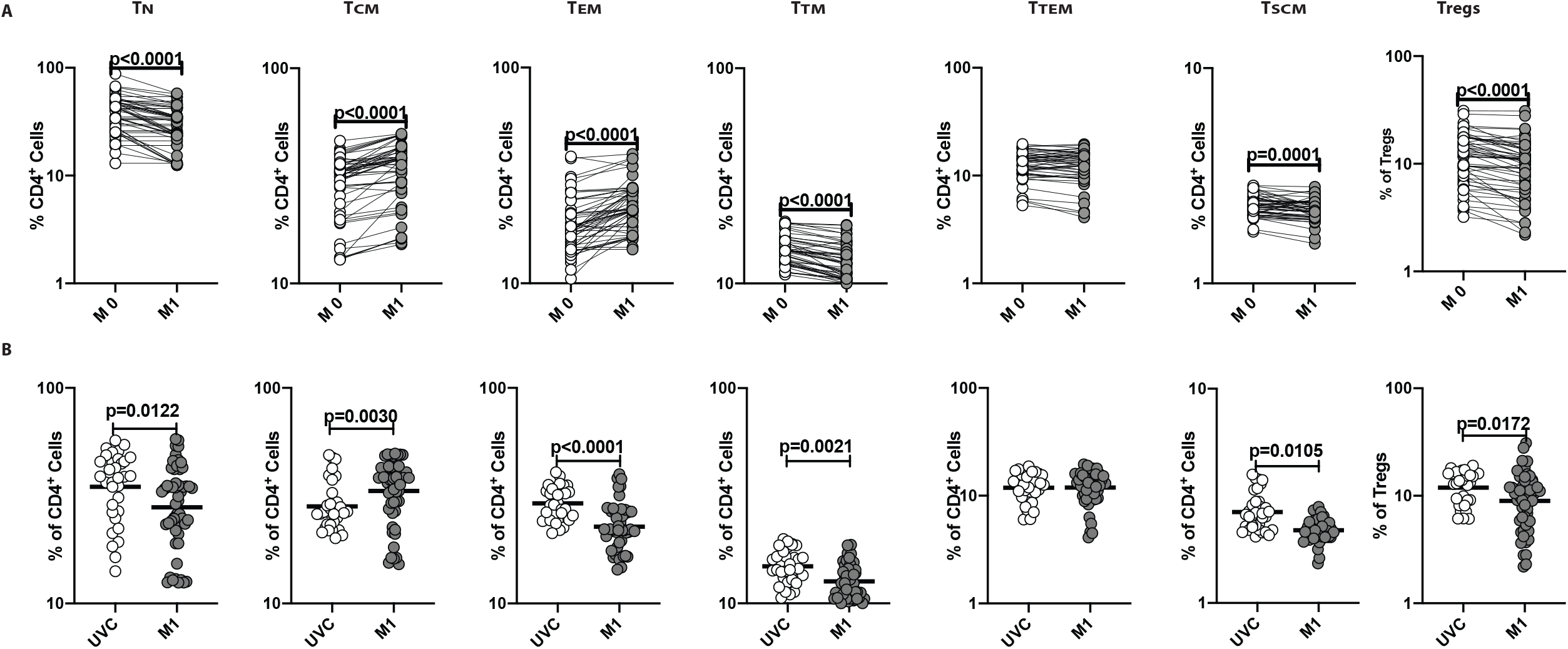
BCG vaccination is associated with altered frequencies of CD4^+^ T cell memory subsets and regulatory cells. (A) Frequencies of CD4^+^ T cell subsets in BCG pre-vaccinated [M0] (n = 54) and month 1 following vaccination [M1] (n = 54). Data are shown as line diagrams with each line representing a single individual. P values were calculated using the Wilcoxon matched pair tests with Holms correction for multiple comparisons. (B) Frequencies of CD4^+^ T cell subsets in BCG unvaccinated (UVC) (n = 32) and post vaccinated [M1] (n = 54) individuals. The data are represented as scatter plots with each circle representing a single individual. P values were calculated using the Mann-Whitney test with Holm’s correction for multiple comparisons.

### BCG vaccination induces enhanced frequencies of central, effector, and terminal effector memory CD8+ T cells and diminished frequencies of naïve, transitional, and stem cell memory CD8+ T cells

To assess the ex vivo phenotype of CD8+ T cell subsets following BCG vaccination, we compared the subsets at baseline or before BCG vaccination (M0) and at month 1 (M1) post-vaccination. As shown in Figure 2A, the frequencies of central, effector, and terminal effector memory CD8+ T cell subsets were significantly enhanced and the frequencies of naïve, transitional, and stem cell memory CD8+ T cells were significantly diminished at M1 compared to M0. Next, we compared the frequencies of CD8+ T cell subsets in post-vaccinated individuals to unvaccinated controls. As shown in Figure 2B, BCG vaccinated individuals exhibited increased frequencies of central memory, effector memory, and terminal effector memory CD8+ T cells and decreased frequencies of naïve, transitional memory, and stem cell memory CD8+ T cells. A representative flow cytometry plot showing the gating strategy for CD8+ T cell subsets is shown in S. Figure 1. Thus, BCG vaccination induces enhanced frequencies of central, effector, and terminal effector memory CD8+ T cells in elderly individuals.

**Figure 2.**
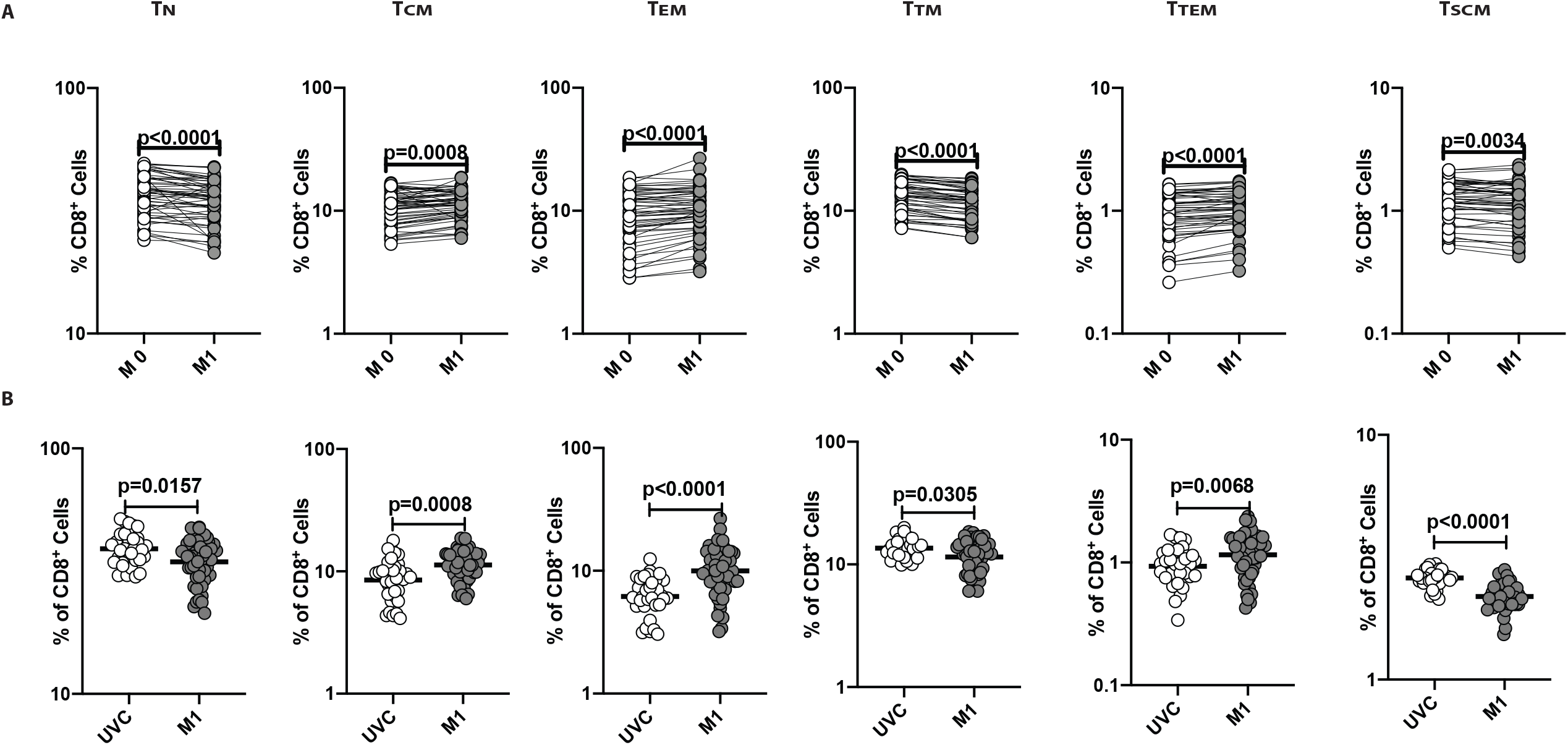
BCG vaccination is associated with altered frequencies of CD8^+^ T cell memory subsets. (A) Frequencies of CD8^+^ T cell subsets in BCG pre-vaccinated [M0] (n = 54) and month 1 following vaccination [M1] (n = 54). Data are as shown line diagrams with each line representing a single individual. P values were calculated using the Wilcoxon matched pair tests with Holms correction for multiple comparisons. (B) Frequencies of CD8^+^ T cell subsets in BCG unvaccinated (UVC) (n = 32) and post vaccinated [M1] (n = 54) individuals. The data are represented as scatter plots with each circle representing a single individual. P values were calculated using the Mann-Whitney test with Holm’s correction for multiple comparisons.

### BCG vaccination induces enhanced frequencies of immature, classical memory and activated memory B cells and plasma cells and diminished frequencies of naïve and atypical memory B cells

To assess the ex vivo phenotype of B cell subsets following BCG vaccination, we compared the subsets at baseline or before BCG vaccination (M0) and at month 1 (M1) post-vaccination. As shown in Figure 3A, the frequencies of immature, classical memory and activated memory B cells and plasma cells were significantly enhanced and the frequencies of naïve and atypical memory B cells were significantly diminished at M1 compared to M0. Next, we compared the frequencies of B cell subsets in post-vaccinated individuals to unvaccinated controls. As shown in Figure 3B, BCG vaccinated individuals exhibited increased frequencies of immature, classical memory and activated memory B cells and plasma cells and decreased frequencies of naïve and atypical memory B cells. A representative flow cytometry plot showing the gating strategy for B cell subsets is shown in S. Figure 2. Thus, BCG vaccination induces enhanced frequencies of immature, classical, and activated memory B cells and plasma cells in elderly individuals.

**Figure 3.**
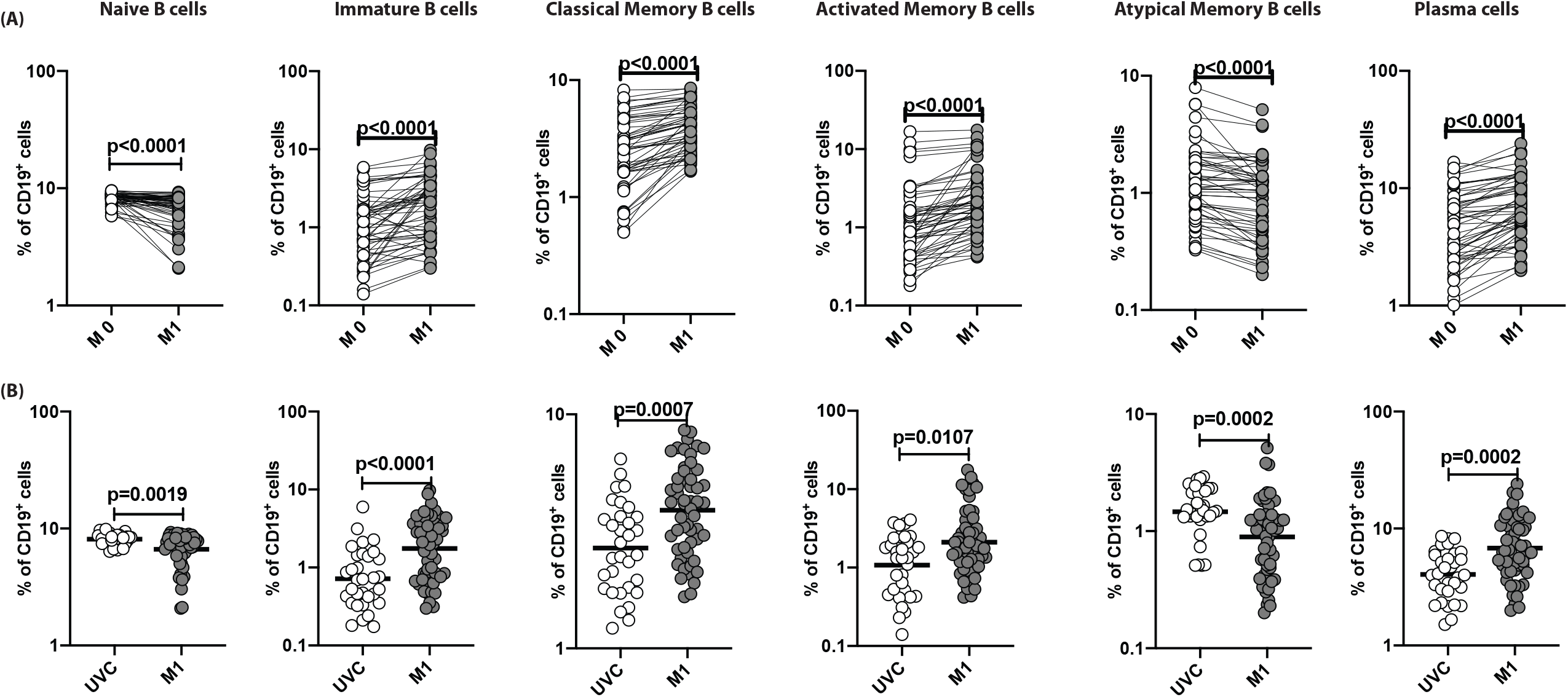
BCG vaccination is associated with altered frequencies of B cells subsets. (A) Frequencies of B cell subsets in BCG pre-vaccinated [M0] (n = 54) and month 1 following vaccination [M1] (n = 54). Data are as shown line diagrams with each line representing a single individual. P values were calculated using the Wilcoxon matched pair tests with Holms correction for multiple comparisons. (B) Frequencies of B cell subsets in BCG unvaccinated (UVC) (n = 32) and post vaccinated [M1] (n = 54) individuals. The data are represented as scatter plots with each circle representing a single individual. P values were calculated using the Mann-Whitney test with Holm’s correction for multiple comparisons.

### BCG vaccination induces enhanced frequencies of myeloid and plasmacytoid DCs but no significant alterations in monocyte subsets

To assess the ex vivo phenotype of monocyte subsets following BCG vaccination, we compared the subsets at baseline or before BCG vaccination (M0) and at month 1 (M1) post-vaccination. As shown in Figure 4A, the frequencies of classical, intermediate, or non-classical monocytes were not significantly altered at M1 compared to M0 in BCG vaccinated individuals. Next, we compared the frequencies of monocyte subsets in post-vaccinated individuals to unvaccinated controls. As shown in Figure 3B, BCG vaccinated individuals exhibited increased frequencies of only intermediate monocytes in comparison to unvaccinated controls. A representative flow cytometry plot showing the gating strategy for monocyte subsets is shown in S. Figure 3. Thus, BCG vaccination does not induce any significant alterations in the frequencies of monocyte subsets in elderly individuals.

**Figure 4.**
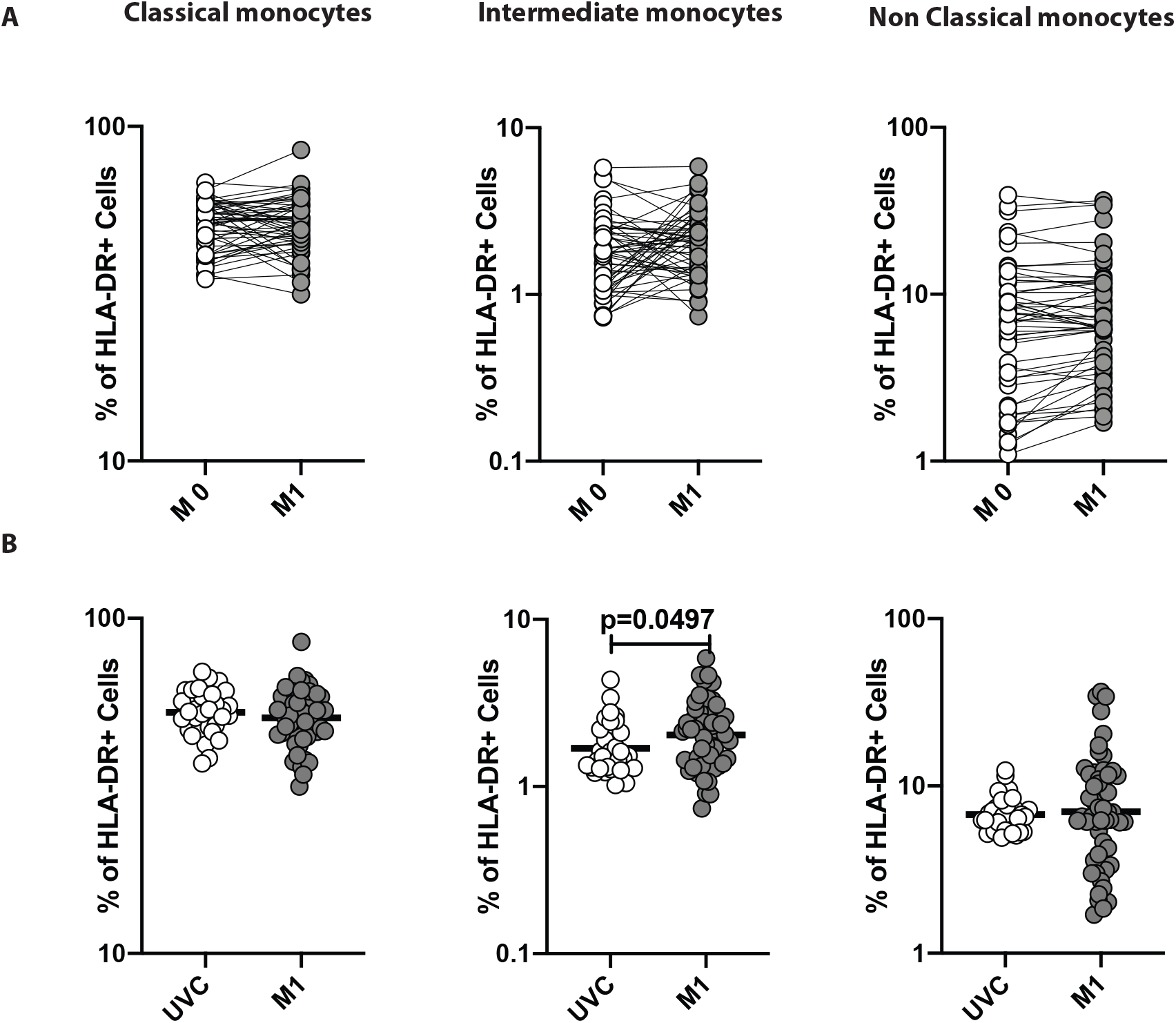
BCG vaccination is not associated with alteration in monocyte subsets. (A) Frequencies of monocyte (classical, intermediate and non-classical) subsets in BCG pre-vaccinated [M0] (n = 54) and month 1 following vaccination [M1] (n = 54). Data are as shown line diagrams with each line representing a single individual. P values were calculated using the Wilcoxon matched pair tests with Holms correction for multiple comparisons. (B) Frequencies of monocyte (classical, intermediate and non-classical) subsets in BCG unvaccinated (UVC) (n = 32) and post vaccinated [M1] (n = 54) individuals. The data are represented as scatter plots with each circle representing a single individual. P values were calculated using the Mann-Whitney test with Holm’s correction for multiple comparisons.

To assess the ex vivo phenotype of DC subsets following BCG vaccination, we compared the subsets at baseline or before BCG vaccination (M0) and at month 1 (M1) post-vaccination. As shown in Figure 5A, the frequencies of myeloid and plasmacytoid DCs were significantly increased at M1 compared to M0 in BCG vaccinated individuals. Next, we compared the frequencies of DC subsets in post-vaccinated individuals to unvaccinated controls. As shown in Figure 5B, BCG vaccinated individuals exhibited increased frequencies of only both myeloid and plasmacytoid DCs in comparison to unvaccinated controls. A representative flow cytometry plot showing the gating strategy for DC subsets is shown in S. Figure 3. Thus, BCG vaccination-induces enhanced frequencies of DC subsets in elderly individuals.

**Figure 5.**
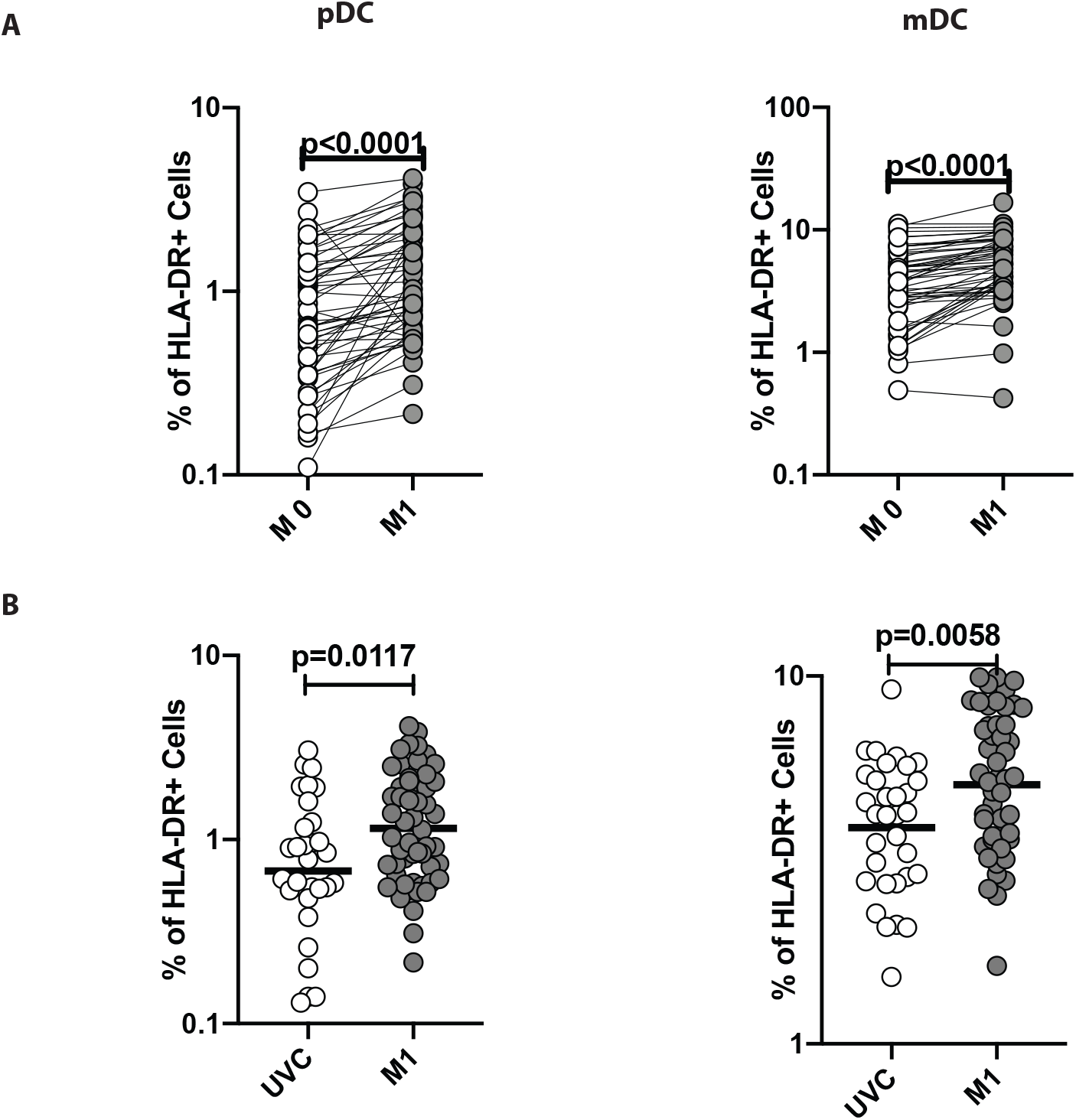
BCG vaccination is associated with heightened frequencies of dendritic cell subsets. (A) Frequencies of dendritic cell (DC) subsets (plasmacytoid DC and myeloid DC) in BCG pre-vaccinated [M0] (n = 54) and month 1 following vaccination [M1] (n = 54). Data are as shown line diagrams with each line representing a single individual. P values were calculated using the Wilcoxon matched pair tests with Holms correction for multiple comparisons. (B) Frequencies of dendritic cells (DC) subsets (plasmacytoid DC and myeloid DC) in BCG unvaccinated (UVC) (n = 32) and post vaccinated [M1] (n = 54) individuals. The data are represented as scatter plots with each circle representing a single individual. P values were calculated using the Mann-Whitney test with Holm’s correction for multiple comparisons.

### BCG vaccination induces increased levels of total IgG subtypes, IgA and IgM

To assess the levels of humoral responses induced by BCG vaccination, we measured IgG subtypes (IgG1, IgG2, IgG3, IgG4), IgA, and IgM in BCG vaccinated individuals at baseline (M0) and month 1 (M1) following BCG vaccination. As shown in Figure 6, BCG vaccination induces significantly increased levels of IgG1, IgG2, IgG3, IgG4, IgA, and IgM at M1 compared to M0. Therefore, BCG is a potent inducer of all antibody isotypes in elderly individuals.

**Figure 6.**
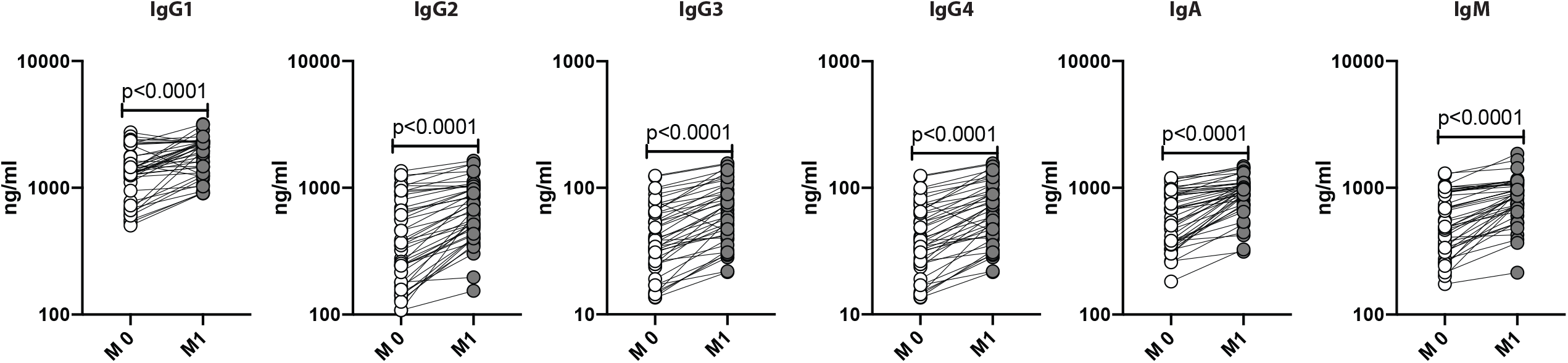
BCG vaccination is associated with heightened frequencies of IgG Isotypes. The plasma levels of IgG isotypes (IgG1, IgG2, IgG3, IgG4), IgA and IgM were measured in BCG pre-vaccinated [M0] (n = 54) and post vaccinated [M1] (n = 54) individuals. The data are represented as scatter plots with each circle representing a single individual. P values were calculated using the Mann-Whitney test with Holm’s correction for multiple comparisons.

## Discussion

Typically, elderly individuals are at high risk of infectious diseases. In the context of COVID-19 disease, elderly individuals are one of the main target groups for morbidity and mortality. Several clinical trials are currently underway to examine the effect of BCG vaccination in protecting health care workers and other individuals from SARS-CoV2 infection or disease (19-22). Very few studies are examining the protective effect of BCG vaccination against SARS-CoV2 in elderly individuals (21). The study to evaluate the effectiveness of the BCG vaccine in reducing morbidity and mortality in elderly individuals in COVID-19 hotspots in India undertaken by the Indian Council of Medical Research is one such study. As part of the study protocol, we examined the immune responses engendered by BCG vaccination in a group of elderly individuals. Previous studies in elderly individuals have shown that BCG vaccination protected against respiratory infections in Indonesia, Japan, and Europe (23-25).

Memory T cells are important in providing vaccine-induced protection against infections in elderly individuals (26). Memory CD4+ T cells induced by vaccination, especially central and effector memory CD4+ T cells, are involved in preventing varicella-zoster reactivation in older individuals (27, 28). Memory CD8+ T cells induced by vaccination, mainly central and effector memory CD8+ T cells, are involved in providing immunity against viral pathogens such as influenza and respiratory syncytial virus (29-32). While other CD4+ and CD8+ memory T cell subsets, including transitional memory, terminal effector memory, and stem cell memory subsets, have been postulated to play a role in infection and cancer (33-35), their exact role in vaccine-elicited human immune responses is still unclear. However, induction of vaccine-induced T cell responses is clearly impaired in elderly individuals (26). One potential mechanism is the expansion of regulatory CD4+ T cells, which are known to down modulate effector T cell responses against pathogens (36).

Our study clearly demonstrates the increased frequencies of both CD4+ and CD8+ central and effector memory T cells. In addition, terminal effector CD8+ T cell memory frequencies are also increased. This is associated with a decrease in the other CD4+ and CD8+ T cell subsets. Therefore, the heightened frequency of the central and effector memory T cell compartment signifies a potential impact on the heterologous immunity and suggests that non-specific or bystander infections are more likely to be protected against in BCG vaccinated individuals. These data fit very well with the recent finding that BCG vaccination in elderly patients was associated with increased time to first infection and protection against viral respiratory pathogens (23). Moreover, BCG vaccination also diminishes the frequency of regulatory T cells and could therefore potentially blunt or ameliorate the regulatory effects of these cells in down modulating protective immune responses.

Most of the current vaccines rely on the induction of protective antibodies through the expansion of memory B cells and plasma cells. Memory B cells can be classified as classical memory, activated memory, and atypical memory B cells (37). While classical and activated memory B cells are important for the production of high affinity and broadly neutralizing antibodies (38), atypical memory B cells are exhausted B cells and fail to engender antibody production (39). Long-lived circulating plasma cells are the major source of vaccine-induced antibody production (40). Thus, our data demonstrate that BCG vaccination is likely to have major effects on the humoral immune response as it induces elevated frequencies of all major subsets of antibody-producing and antibody competent B cells. Our data also supports another mechanism for the off-target or non – specific effect of BCG vaccination, which is its ability to cause expansion of memory B cells and plasma cells. Indeed, the examination of different antibody isotypes reveals the presence of enhanced levels of IgG1, IgG2, IgG3, IgG4, IgA and IgM, indicating that humoral responses are significantly induced by BCG vaccination.

BCG vaccination is known to induce trained immunity or innate memory, mainly by inducing epigenetic modification of monocytes and enhancing their ability to mount cytokine responses to secondary pathogens (18). Our data show that BCG vaccination did not significantly modulate the frequencies of monocyte subsets in vaccinated individuals. Thus, the effect of BCG on monocyte responses is unlikely to involve monocyte subset alterations. Activating the adaptive immune response requires the presentation of antigens to T cells by DC and macrophages (41). Delays in activation of DC could often result in a delay in the induction of the adaptive immune response to pathogens. Thus, methods to either increase the frequencies of DCs or to activate DCs would improve the protective efficacy of vaccines (42). Our data showing elevated frequencies of both mDC and pDC thus clearly illustrate an important effect of BCG vaccination in enhancing the innate immune response to both specific and non-specific pathogens (possibly) in elderly individuals. Moreover, while mDC is mainly involved in the process of antigen – presentation (43), pDC is the main source of Type 1 interferons in the host (44). Thus, it is potentially likely that increased frequencies of pDC might heighten the propensity of pDC to mount Type I IFN responses against pathogens encountered.

In summary, our study highlights the effect of BCG vaccination in modulating the frequencies of both innate and adaptive immune cell subsets. Our study also reveals an effect of BCG in inducing heightened total antibody levels. Although our study did not examine the functional effects of these changes in the immune system, our data nevertheless reveal an important role for BCG vaccination in boosting immune responses in the elderly population. Whether this translates to improved protective immunity to non – specific infections like SARS-CoV2 remains to be determined.

## Materials and Methods

### Ethics statement

The study was approved by the Ethics Committees of NIRT (NIRT-INo:2020010). Informed written consent was obtained from all participants. The study is part of the clinical study entitled, Study to evaluate the effectiveness of the BCG vaccine in reducing morbidity and mortality in elderly individuals in COVID-19 hotspots in India (NCT04475302).

### Study Population

Elderly individuals, between 60 - 80 years of age, residing in hotspots for SARS-Cov2 infection were included in the study between July 2020 and September 2020 in Chennai, India after obtaining informed written consent from the study participants.. Elderly population positive for SARS-Cov2 infection by either antibody (serology) or PCR test; HIV infected or individuals with malignancy or on immunosuppressive drugs or transplant recipient and those on dialysis or anti-psychiatric medications or hypersensitivity to vaccinations were not included in the study. Also, those who were diagnosed with tuberculosis (TB) in the previous 6-months or were currently on anti-TB treatment were not included in the study.

54 participants received a single dose of BCG vaccine (Freeze-dried) manufactured by Serum Institute of India, Pune. The adult dose of BCG vaccine was 0.1 mL injected intradermally over the distal insertion of the deltoid muscle onto the left humerus (approximately one third down the left upper arm). In case of a previous vaccination scar, or presence of ulcer/injury or tattoo on the left upper arm, vaccination was given in the right upper arm. 32 elderly individuals from the same hotspot area were not vaccinated and were considered as controls. Blood was drawn from the vaccinated participants at baseline (before vaccination) and 1 month following vaccination. Blood was drawn from the controls only at baseline

### Ex vivo analysis

All antibodies used in the study were from BD Biosciences (San Jose, CA), BD Pharmingen (San Diego, CA), eBioscience (San Diego, CA), or R&D Systems (Minneapolis, MN). Whole blood was used for ex vivo phenotyping and it was performed on all 87 individuals. Briefly, to 250μl aliquots of whole blood a cocktail of monoclonal antibodies specific for various immune cell types was added. T cell phenotyping was performed using antibodies directed against CD45-Peridinin chlorophyll protein (PerCP), CD3- phycoerythrin (PE) Cy7, CD-8 AmCyan, CD28- allophycocyanin (APC) H7, CD45RA-Pacific Blue, and CCR7-FITC and CD95- PE. Naive cells were classified as CD45RA^+^ CCR7^+^ CD95^-^ CD28^+^, central memory cells (TCM) as CD45RA^-^ CCR7^+^ CD95^+^ CD28^+^, effector memory cells (TEM) as CD45RA^-^CCR7^-^ CD95^+^ CD28, Terminal effector memory cells (TTEM) as CD45RA^-^ CCR7^-^ CD95^+^ CD28^-^, stem cell memory (TSCM) as CD45RA^+^ CCR7^+^ CD95^+^ CD28^+^ and transitional memory cells (TTM) as CD45RA^+^ CCR7^-^ CD95+ CD28+ (45). Regulatory T cell phenotyping was performed using CD3 APC-Cy-7, CD4 Amycyn, CD8 PerCP, CD25 APC, CD127 FITC, Foxp3 PE and regulatory T cells were classified as CD4^+^ CD25^+^ Foxp3^+^ CD127dim (45). B cell phenotyping was performed using antibodies directed against CD45-PerCP, CD19-Pacific Blue, CD27-APC-Cy7, CD21-FITC, CD20-PE and CD10-APC. Naive B cells were classified as CD45^+^ CD19^+^ CD21^+^ CD27-; classical memory B cells as CD45^+^ CD19^+^ CD21^+^ CD27^+^; activated memory B cells as CD45^+^ CD19^+^ CD21- CD27^+^; atypical memory B cells as CD45^+^ CD19^+^ CD21-CD27-; immature B cells as CD45^+^ CD19^+^ CD21^+^ CD10^+^; and plasma cells as CD45^+^ CD19^+^ CD21- CD20- (39). Phenotyping of DC was performed using antibodies directed against HLA- DR, lineage cocktail (CD3, CD14, CD16, CD19, CD20, CD56) FITC CD123-PE (clone 9F5; BD) CD11c- APC. Plasmacytoid DCs were classified as (Lin– HLA-DR^+^ CD123^+^) and myeloid DCs (Lin– HLA- DR^+^ CD11c^+^). Monocyte phenotyping was performed using antibodies directed against CD45- PerCP, CD14-Pacific Blue, HLA-DR-PE-Cy7 (clone L243; BD), and CD16-APC- Cy7. Classical monocytes were classified as CD45^+^ HLA-DR^+^ CD14^hi^ CD16-; intermediate monocytes as CD45+ HLA-DR+ CD14^hi^ CD16^dim^ and non-classical monocytes were classified as CD45+ HLA- DR+CD14^dim^CD16^hi^. Following 30 min of incubation at room temperature erythrocytes were lysed using 2 ml of FACS lysing solution (BD Biosciences Pharmingen), and cells were washed twice with 2 ml of 1XPBS and suspended in 200 μl of PBS (Lonza, Walkersville, MD). Eight- color flow cytometry was performed on a FACS Canto II flow cytometer with FACSDIVA software, version 6 (Becton Dickinson). The gating was set by forward and side scatter, and 100 000 gated events were acquired. Data were collected and analyzed using FLOW JO software (TreeStar, Ashland, OR). Leukocytes were gated using CD45 expression versus side scatter (46, 47).

### Antibody Isotyping

Circulating plasma levels of IgG1, IgG2, IgG3, IgG4, IgA, and IgM were measured using Human Isotyping Assays (Bio-Rad-Bio-Plex ProTM) ELISA kits according to manufacturer’s instruction.

### Statistical analysis

Geometric means (GM) were used for measurements of central tendency. Wilcoxon signed-rank test was used to compare frequencies of immune subsets in the BCG vaccinated group at month 0 (M0) and month 1 (M1). Statistically significant differences between unvaccinated and BCG vaccinate M1 groups were analyzed using the Mann-Whitney test. Analyses were performed using Graph-Pad PRISM Version 8.0.

## Data Availability

All the reported data are available within the manuscript

## Author Contributions

Designed the study (S.B., C.P); conducted experiments (N.P.K., R.A., A.N., N.S., R.M.R, V.V); acquired data (N.P.K., R.A., A.N.); analyzed data (N.P.K., R.A); contributed reagents and also revised subsequent drafts of the manuscript (S.B., C.P); responsible for the enrolment of the participants and also contributed to acquisition and interpretation of clinical data (C.P., B.P.K., B.J., D.K., S.T); wrote the manuscript (S.B., N.P.K., C.P). All authors read and approved the final manuscript.

## Acknowledgments

We thank the Director of the ICMR-NIRT, and staff of the Department of Clinical Research, NIRT. We thank the data entry operators Mr. Jaiganesh and Mr. Vigneshwaran, and also all the staff members of the ICER department for the timely help.

## Data and materials availability

All the reported data are available within the manuscript

---

**Supplementary Figure 1.**
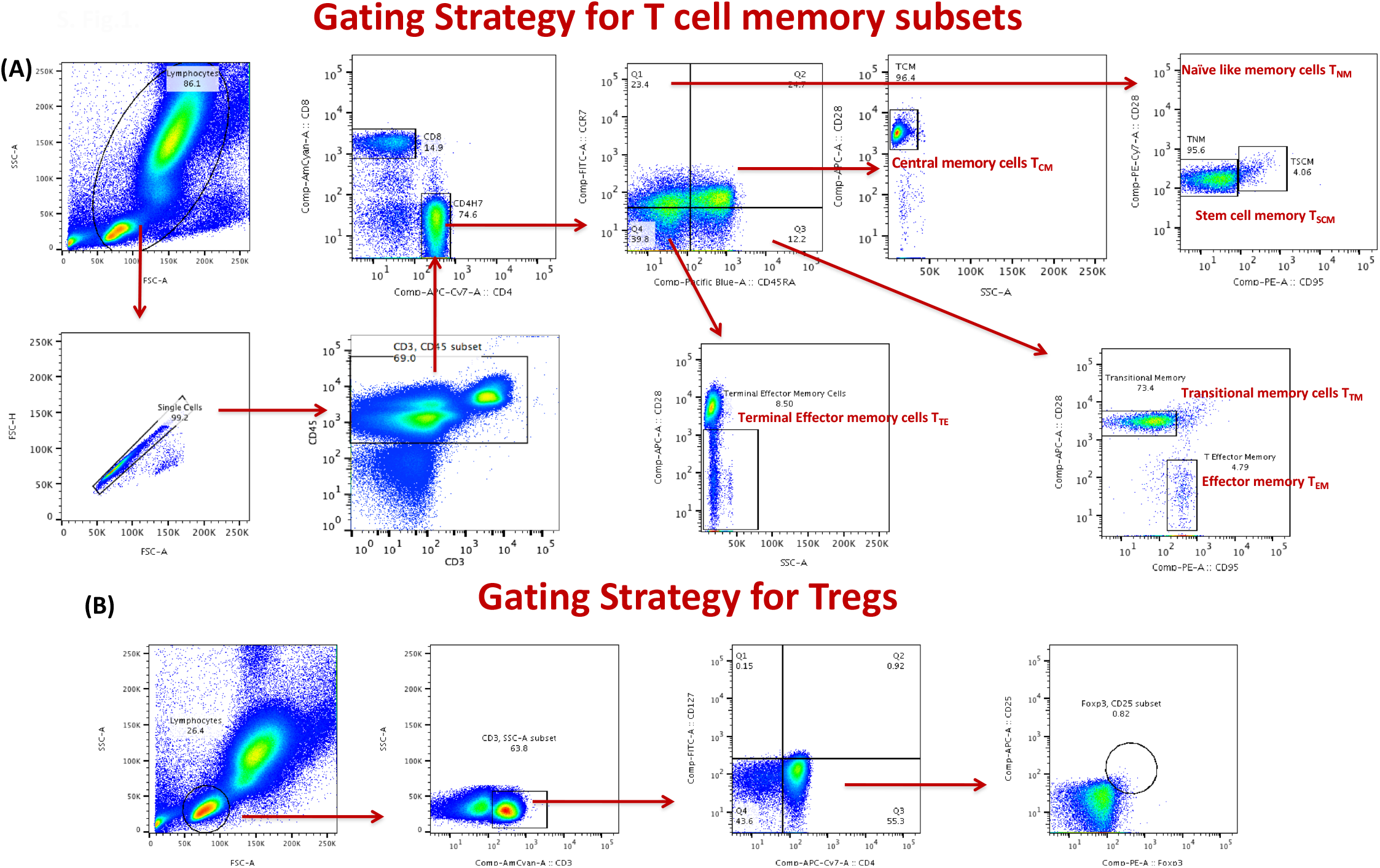
Gating strategy for CD4+ and CD8+ T cell subsets. (A) A representative flow cytometry plot from an BCG vaccinated individual at month 0 showing the gating strategy for Naïve cells (T_N_) were classified as CD45RA^+^ CCR7^+^ CD95^-^ CD28^+^, central memory cells (T_CM_) as CD45RA^-^ CCR7^+^ CD95^+^ CD28^+^, effector memory cells (T_EM_) as CD45RA^-^CCR7^-^ CD95^+^ CD28^-^, Terminal effector memory cells (T_E_) as CD45RA^-^ CCR7^-^ CD95^+^ CD28^-^, stem cell memory (T_SCM_) as CD45RA^+^ CCR7^+^ CD95^+^ CD28^+^ and transitional memory cells (T_TM_) as CD45RA^+^ CCR7^-^ CD95^+^ CD28^+^. (B) Regulatory T cells were classified as CD4^+^ CD25^+^ Foxp3^+^ CD127dim

**Supplementary Figure 2.**
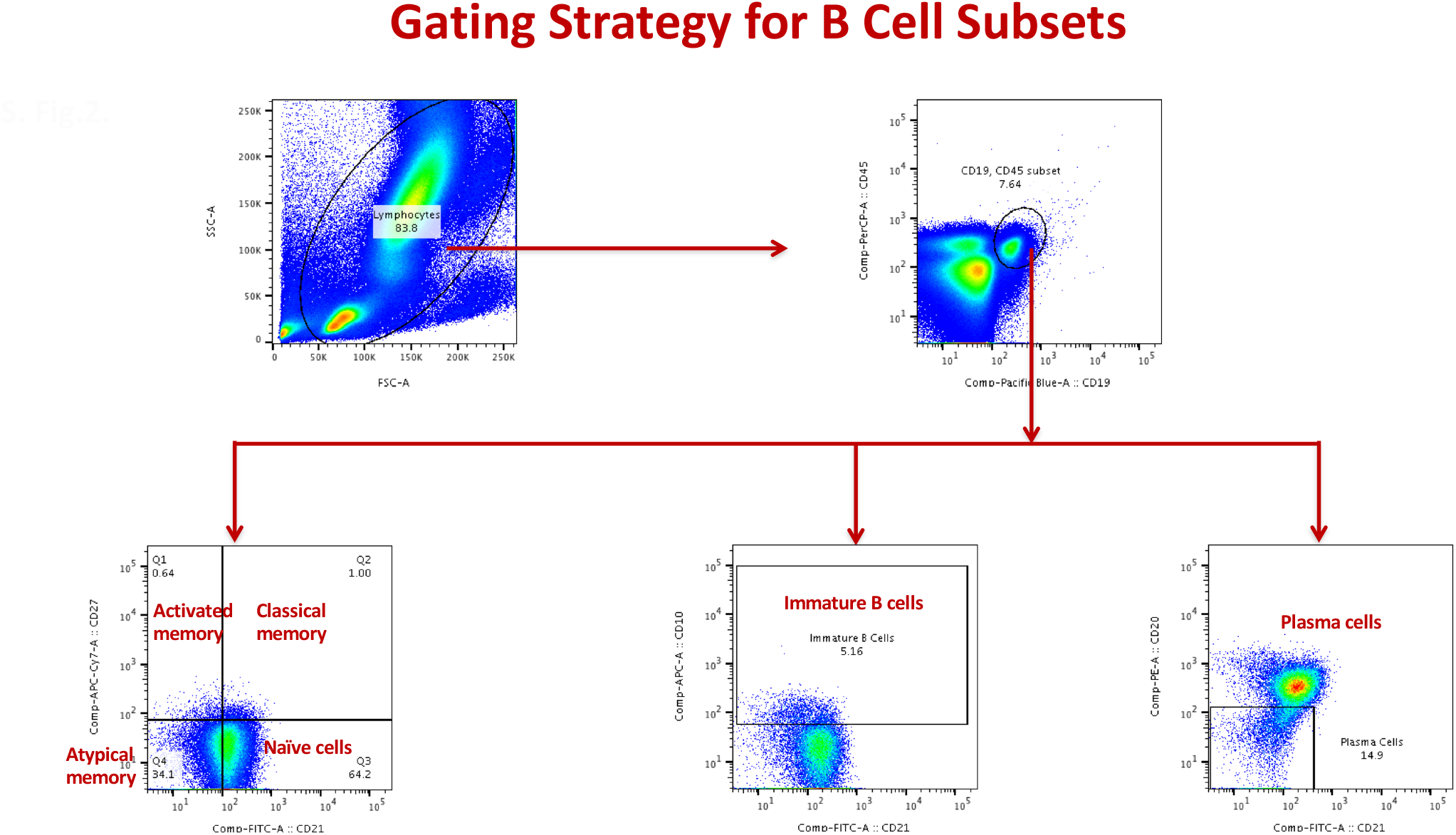
Gating strategy for B cell subsets. (A) A representative flow cytometry plot from an BCG vaccinated individual at month 0 showing the gating strategy for naïve, immature, classical memory (CM), activated memory (AM), Atypical memory (ATM), immature and plasma cells from CD45+ CD19+ cells. Naïve cells were classified as CD21+ CD27-; classical memory (CM) cells as CD21+ CD27+; activated memory (AM) cells as CD21- CD27+; Atypical memory (ATM) cell as CD21- CD27-; immature B cells as CD21+ CD10+; and plasma cells as CD21- CD27-.

**Supplementary Figure 3.**
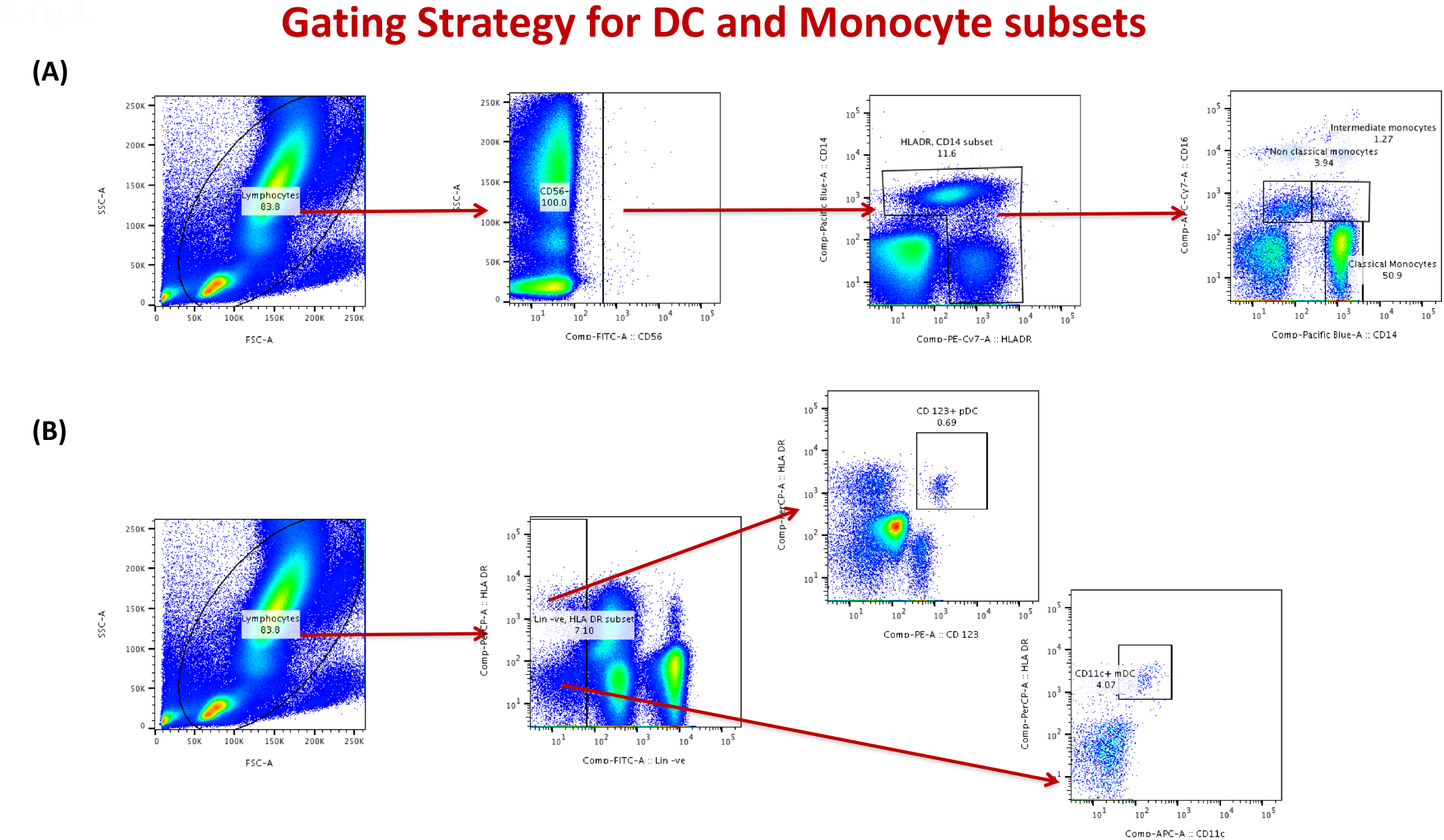
Gating strategy for DC and monocyte subsets. (A) A representative flow cytometry plot from an BCG vaccinated individual at month 0 showing the gating strategy for plasmacytoid (pDC) and myeloid DCs (mDC) Plasmacytoid DC were classified as (Lin^−^ HLA-DR^+^ CD123^+^) and myeloid DCs as (Lin^−^ HLA-DR^+^ CD11c^+^). (B) A representative flow cytometry plot showing the gating strategy for estimation of monocyte subsets. Classical monocytes were classified as CD45^+^ HLA-DR^+^ CD14^hi^CD16^−^; intermediate monocytes as CD45^+^ HLA-DR^+^ CD14^hi^ CD16^dim^ and non-classical monocytes were classified as CD45^+^HLADR^+^ CD14^dim^CD16^hi^.

