## Supplementary material for "BCG vaccination induces enhanced frequencies of memory T and B cells and dendritic cell subsets in elderly individuals": S. Fig.1

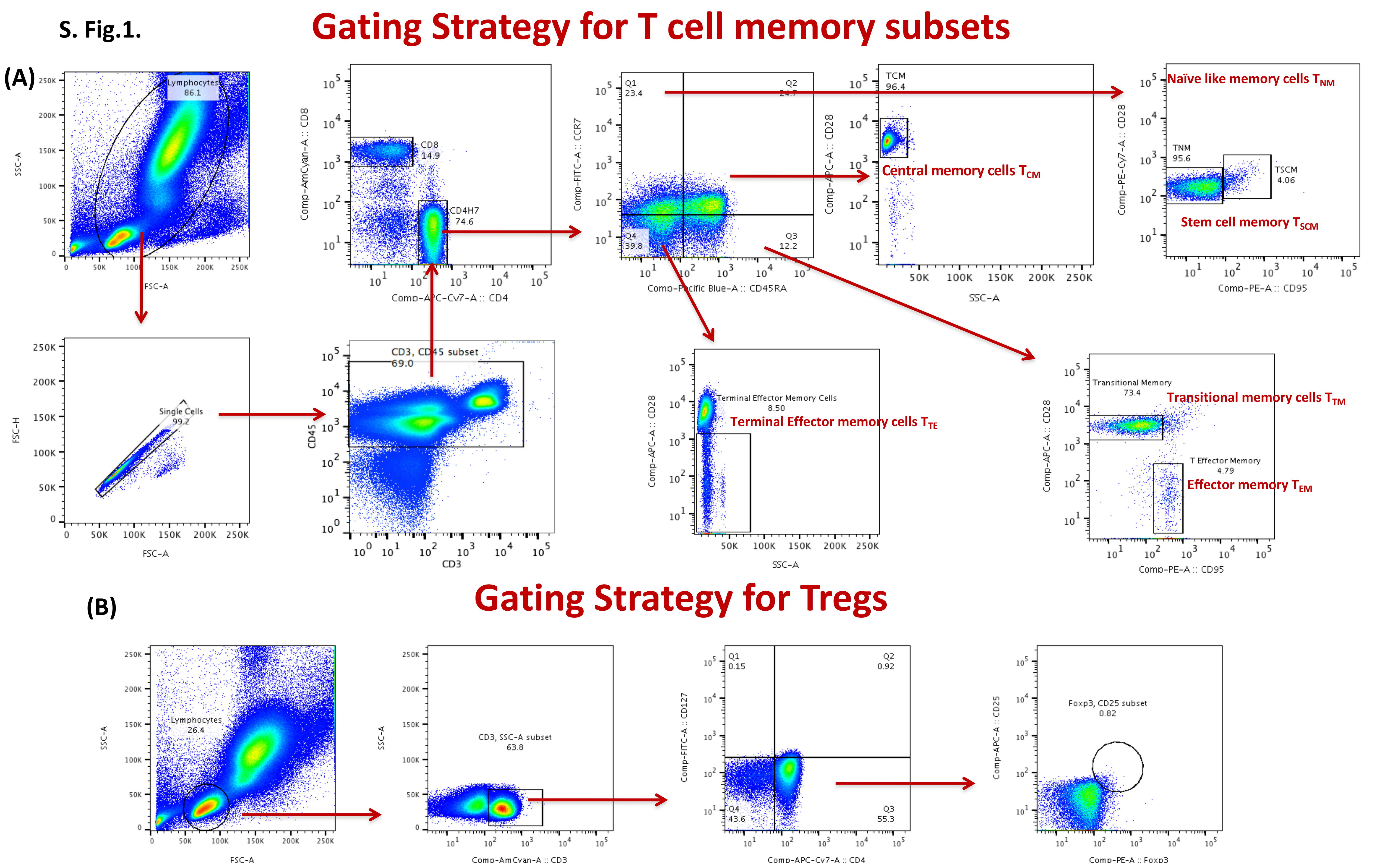


**Supplementary Figure 1. Gating strategy for CD4+ and CD8+ T cell subsets**. (A) A representative flow cytometry plot from an BCG vaccinated individual at month 0 showing the gating strategy for Naïve cells ($T_N$) were classified as CD45RA$^+$ CCR7$^+$ CD95$^-$ CD28$^+$, central memory cells ($T_{CM}$) as CD45RA$^-$ CCR7$^+$ CD95$^+$ CD28$^+$, effector memory cells ($T_{EM}$) as CD45RA$^-$CCR7$^-$ CD95$^+$ CD28$^-$, Terminal effector memory cells ($T_E$) as CD45RA$^-$ CCR7$^-$ CD95$^+$ CD28$^-$, stem cell memory ($T_{SCM}$) as CD45RA$^+$ CCR7$^+$ CD95$^+$ CD28$^+$ and transitional memory cells ($T_{TM}$) as CD45RA$^+$ CCR7$^-$ CD95$^+$ CD28$^+$. (B) Regulatory T cells were classified as CD4$^+$ CD25$^+$ Foxp3$^+$ CD127dim
