## Supplementary material for "BCG vaccination induces enhanced frequencies of memory T and B cells and dendritic cell subsets in elderly individuals": S.Fig.2

### Gating Strategy for B Cell Subsets

**S. Fig.2.**

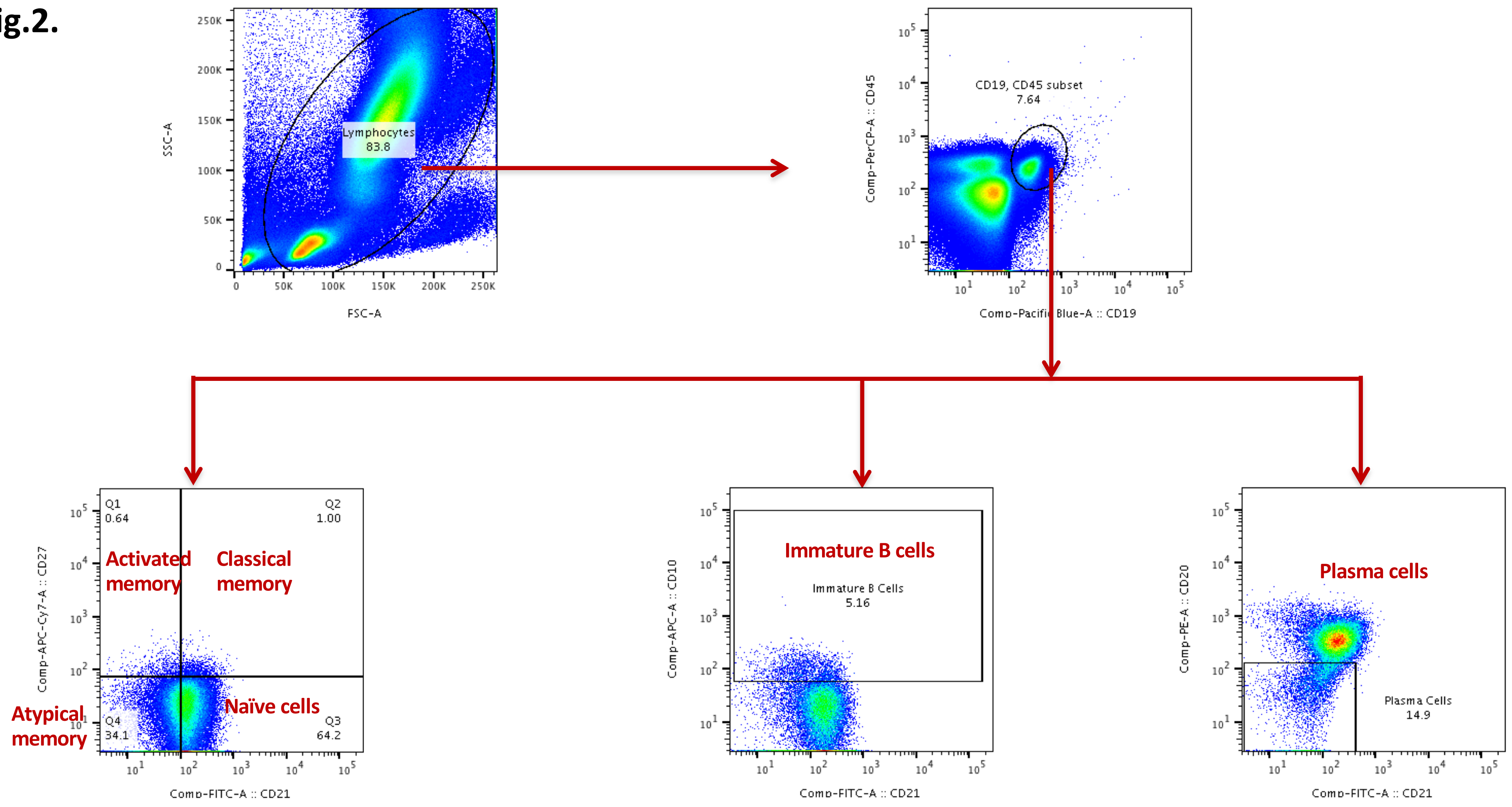


**Supplementary Figure 2. Gating strategy for B cell subsets.** (A) A representative flow cytometry plot from an BCG vaccinated individual at month 0 showing the gating strategy for naïve, immature, classical memory (CM), activated memory (AM), Atypical memory (ATM), immature and plasma cells from CD45+ CD19+ cells. Naïve cells were classified as CD21+ CD27-; classical memory (CM) cells as CD21+ CD27+; activated memory (AM) cells as CD21- CD27+; Atypical memory (ATM) cell as CD21- CD27-; immature B cells as CD21+ CD10+; and plasma cells as CD21- CD27-.
