## Supplementary material for "BCG vaccination induces enhanced frequencies of memory T and B cells and dendritic cell subsets in elderly individuals": S.Fig.3

### Gating Strategy for DC and Monocyte subsets

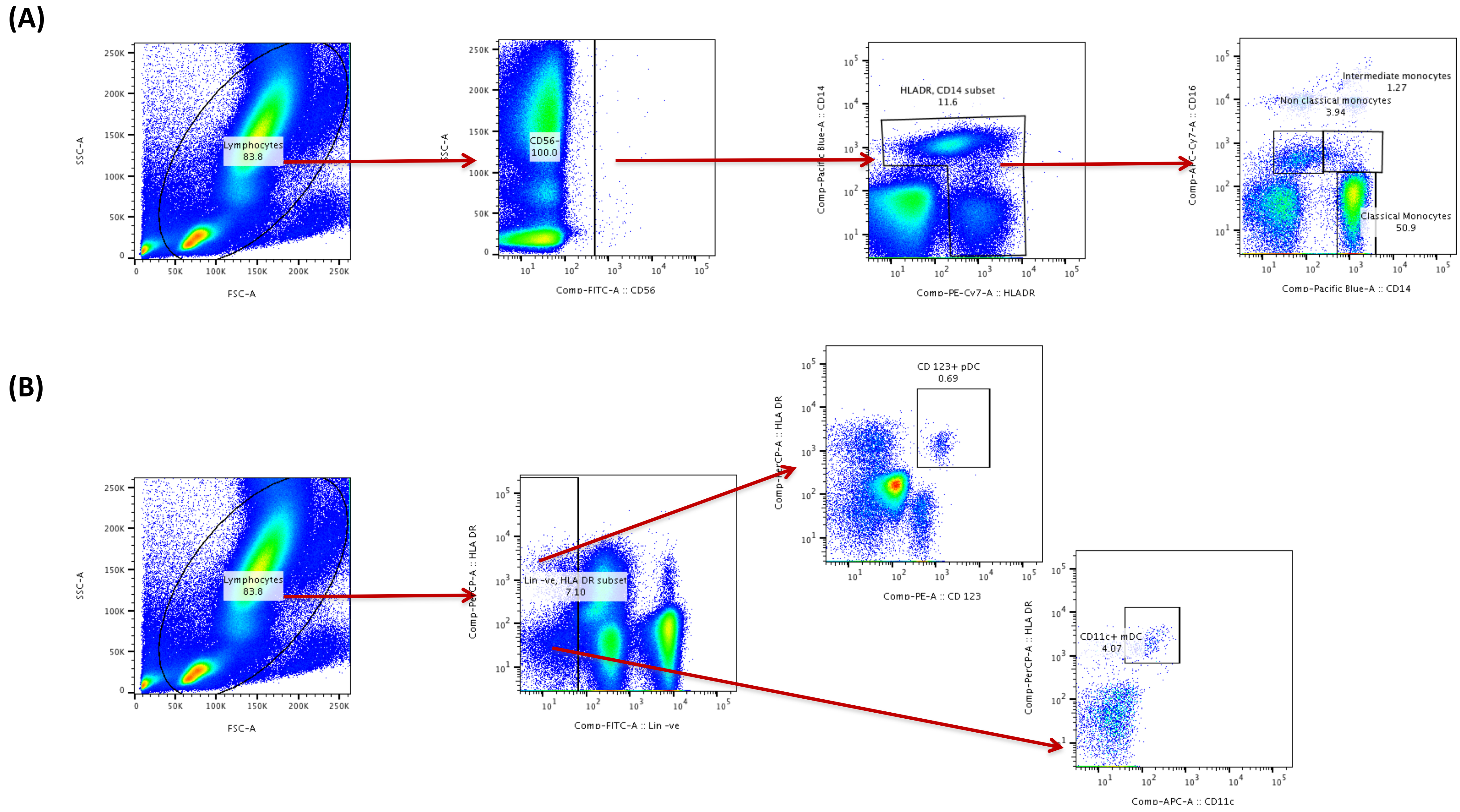


**Supplementary Figure 3. Gating strategy for DC and monocyte subsets** (A) A representative flow cytometry plot from an BCG vaccinated individual at month 0 showing the gating strategy for plasmacytoid (pDC) and myeloid DCs (mDC) Plasmacytoid DC were classified as (Lin$^{-}$ HLA-DR$^{+}$ CD123$^{+}$) and myeloid DCs as (Lin$^{-}$ HLA-DR$^{+}$ CD11c$^{+}$). (B) A representative flow cytometry plot showing the gating strategy for estimation of monocyte subsets. Classical monocytes were classified as CD45$^{+}$ HLA-DR$^{+}$ CD14$^{hi}$CD16$^{-}$; intermediate monocytes as CD45$^{+}$ HLA-DR$^{+}$ CD14$^{hi}$ CD16$^{dim}$ and non-classical monocytes were classified as CD45$^{+}$HLADR$^{+}$ CD14$^{dim}$CD16$^{hi}$.
